# Practical application of CO_2_ as an indicator regarding the risk of infection

**DOI:** 10.1101/2022.07.14.22277631

**Authors:** Anne Hartmann, Yunus Emre Cetin, Petra Gastmeier, Martin Kriegel

## Abstract

The air quality of enclosed spaces has attracted great attention due to the ongoing Covid-19 pandemic. The infection risk in these spaces can be estimated for various scenarios with different methods so the important parameters and effective infection prevention measures can be compared. Previous studies showed that indoor CO_2_ concentration could be considered a surrogate for infection risk. In this regard, a generic relation can be established between the CO_2_ levels and infection probability. Based on this consideration, some practical evaluations between CO_2_ concentration and infection risk are conducted in this study. The effect of mask efficiency, viral emission rate, and duration of exposure are also included in the assessments. It is shown that continuous CO_2_ monitoring can be helpful in the evaluation of possible preventive measures. Findings are expected to contribute to the understanding of the simple parameters related to the infection risk.

## Introduction

The ongoing COVID-19 pandemic has raised global interest on indoor air quality (IAQ) and ventilation, particularly in public spaces [2, 3]. The air quality of these spaces can be assessed with various parameters. Among these, the CO_2_ concentration is considered an important indicator of adequate ventilation [4, 5, 6]. Today, indoor CO_2_ concentrations can be monitored easily by various low-cost sensors with reasonable accuracy [6, 7, 8].

The idea of monitoring CO_2_ concentration as an indication of efficient ventilation is based on the first discussions regarding ventilation in 1858 [9, 10]. In enclosed environments, high CO_2_ concentrations can be reached due to occupants who are mostly the main source of gaseous emissions. Recent findings point out that high CO_2_ levels in indoor spaces may cause detrimental outcomes on health and cognitive function [11, 12]. In relation to these concerns, some recommendations can be found from ASHRAE 62.1 [13], ISO 17772 [14], EN 16798 [15] and UBA [16, 17] regarding the maximum limits of indoor CO_2_ concentrations. Related recommendations were generally calculated based on the minimum ventilation rates specified in these standards. So far, the recommended CO_2_ values were often considered in regulating demand-controlled ventilation, especially for energy-efficient systems.

In addition to CO_2_, viral aerosols can also be emitted during exhalation by occupants. However, detecting and measuring these virus-laden aerosols is not practical, in contrast to CO_2_ measurement [18]. Since the infection risk is directly related to the rebreathed air, indoor CO_2_ levels can be considered as a proxy of a possible infection risk [19, 20, 21, 22]. In this context, a correlation can be established between the indoor CO_2_ concentration and infection probability.

There are different mathematical models used to evaluate the infection probability in indoor premises. One of these models is the well-known Wells-Riley model which is based on the quantum concept [23, 24, 25]. Several modifications of the Wells-Riley model (including the CO_2_ consideration) were developed to calculate the infection risk under varying circumstances. Rudnick and Milton [22] developed a CO_2_-based infection risk evaluation method that determines the rebreathed fraction using CO_2_ concentration as a marker for exhaled breath exposure. By using this method, infection probability of different diseases including measles, influenza, and rhinovirus was assessed. They also showed that the infection risk can be calculated using CO_2_-based evaluation without steady-state concentration assumption and measuring the outdoor air supply rate. Issarow et al. [26] established a similar mathematical model that predicts the risk of airborne infectious diseases, under steady-state and non-steady-state conditions by monitoring exhaled air from infectors. They used the rebreathed air rate concept to directly determine the average volume fraction of exhaled air in a given space. Hartmann and Kriegel [27] suggested a method to assess the infection risk in enclosed environments based on CO_2_ concentration. It was shown that CO_2_ is a good indicator of the efficiency of the ventilation system to eliminate respiratory viruses and is related to the outdoor air supply. Burridge et al. [28] presented a method to determine the relative risk of airborne transmission that depends on CO_2_ data and occupancy levels within an indoor space. It was shown that well-ventilated rooms are unlikely to contribute to airborne infection, while moderate changes in the boundary conditions or new variants may worsen the infection risk. Peng and Jimenez [29] derived analytical expressions of CO_2_-based risk proxies and applied them to different indoor cases. It was disclosed that the relative infection risk can be estimated with CO_2_ concentration and protection can be provided by keeping low CO_2_ rates by ventilation.

In this study, a risk assessment model offered previously by Kriegel et al. [1] is modified considering the relation between infection probability and CO_2_ concentration. In this respect, different cases are scrutinized, and practical outputs are presented in detail. Results are expected to contribute to comprehension of the indoor infection risk and effective parameters on it based on the CO_2_ concentration.

## Material and methods

In the course of a transmission of pathogens, two different categories of risk have to be separated. First, the risk of one specific person in the room to get infected has to be considered. This individual risk can also be seen as an indicator for the percentage of persons in a given situation getting infected. Secondly, the number of persons to which the infection is transferred to by one index case is important to make statements regarding the progress of the pandemic. In case one infected person infects one other person, the number of infected persons remain constant, in case of zero persons it declines and in case of more persons it increases. In the aforementioned model (Kriegel et al [1]) for the situational predicted attack rate (PAR_s_), the percentage of persons within one room/situation getting infected, the virus related factor is expressed as the viral emission rate S_v_ divided by the critical dose N_0_. Since this factor is often not known, especially in the beginning of a pandemic, it will be expressed as the combined factor V_F_ in this study. In addition, a room-related factor (C_R_), the inhalation flow rate of the susceptible persons (Q_b,in_) and the mask efficiency (f_M_) are considered. The complete equation can be seen in equation (1).

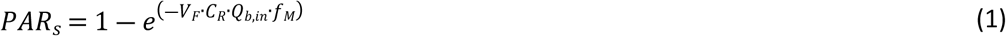

Ideal mixing of the room air is considered for this equation. A first approach to implement different concentrations within the room can be the ventilation effectiveness regarding the German technical norm DIN EN 16798-3:2017-11 [30]. The ventilation effectiveness is defined as the ratio of the difference in concentration between exhaust air (C_e_) and supply air (C_s_) and the difference in concentration between indoor air (C_i_) and supply air. The equation can be seen in (2).

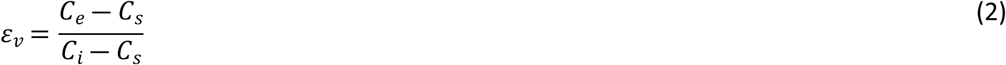

In case of virus laden particles, it is assumed that the outdoor air concentration of these particles is nearly 0 and therefore, if no recirculation or recirculation with high efficient particulate air (HEPA) filter is used, the supply air concentration is 0 as well. Equation (2) can then be transferred into the ratio of exhaust air concentration to indoor air concentration (3).

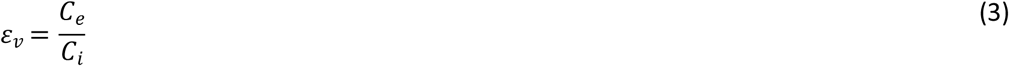

The concentration in the breathing zone of a person is therefore by the ventilation effectiveness higher than the average room concentration used in equation (1), wherefore equation (4) can be set up.

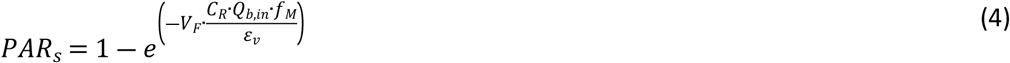

The local ventilation effectiveness can be measured or simulated, but depends on many boundary conditions (e.g. distribution of heat sources (persons) in the room, position of the particle source, …), whereas for a general approach typical values can be used. VDI 3804:2009-03 [31] (VDI means Verein Deutscher Ingenieure, translation: The Association of German Engineers) offers values for situations which deviate from ideal mixing ventilation:

- ideal mixing ventilation: *ε*_*v*_ = 1,0
- mixture of mixing and displacement ventilation: *ε*_*v*_ ≈ 1,2
- displacement ventilation: *ε*_*v*_ ≈ 2,0

To calculate the room-related factor (C_R_) regarding Kriegel et al. [1] the supply air volume flow is necessary. The equations can be seen in (5) for the steady and in (6) for the unsteady situation. It implements the time of stay (t), the virus free supply air volume flow (Q) and the decay rate (λ_g_, consisting of air change, sedimentation and inactivation).

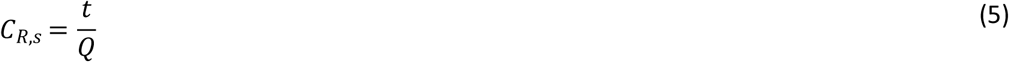

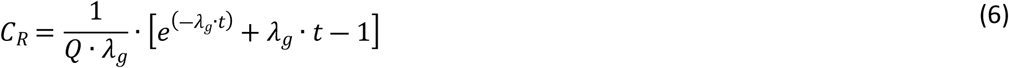

In cases with mechanical ventilation systems, the supply air volume flow is in general designed to reach a certain level, but might still be unknown to the room users. It is also hard to find in rooms, which are ventilated by windows. In cases of window ventilation, the supply air volume flow depends on the outdoor conditions (especially temperature and wind) and can therefore not be assumed to be constant. For a known or measured outdoor CO_2_-concentration (C_ODA_), the supply air volume flow per person only depends on the activity of the persons and can be calculated from the CO_2_-concentration in the room (C_CO2_). This method can be used for window ventilation as well as mechanically ventilated rooms with unknown supply air volume flow. The activity is thereby expressed as the breathing volume flow (Q_b_). The average CO_2_-concentration in the exhaled breath of a person is 40,000 ppm.

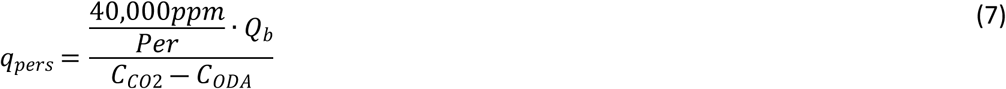

The current limits regarding CO_2_-concentrations e.g. 1000 ppm by Pettenkofer [32] only consider the air quality, but lack an important piece of information, the time of exposure. In a study of Good et al. [33] the emission of CO_2_ was correlated to the emission of particles, which may, in case of an infection carry virus. They showed that an increase in the CO_2_ emission is correlated with an increased particle emission rate, which is for example based on the age and sex of the emitting person. So the CO_2_-emission of the room users is a helpful indicator regarding the risk of infection, but a CO_2_-threshold alone is not useful in description of ventilation regarding infection prevention, but the dose has to be used.

For a practical application of these considerations, three different cases can be considered.

1. A CO_2_-concentration is measured after a certain time of use without a significant change in boundary conditions. The situation in the room can be assumed to be quasi-steady.
2. A CO_2_ threshold is looked for to limit the risk of infection to a certain level.
3. The CO_2_-concentration in the room is constantly monitored and the risk of infection over the time of stay shall be estimated.

### 1. Quasi-steady assumption

With the assumption that critical situations either occur in small rooms or after a long time of stay, a quasi - steady state situation can be assumed. In case of an unsteady situation, this may overestimate the risk. In Kriegel et al. [1] a simplification for equation (4) is derived (see equation (8)).

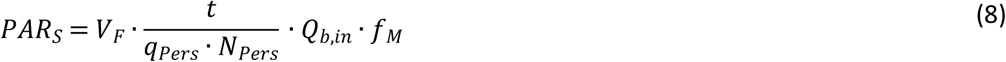

In addition to the steady state, this simplification also assumes that in rooms with intensive ventilation, the discharge of virus laden particles resulting from the air flow will be much higher than the discharge because of sedimentation or inactivation of the potentially virus laden particles, whereas sedimentation and inactivation can be neglected.

From an epidemiological point of view, it should be aimed for that an infected person does not infect more than one additional person. Because only one situation is considered in this investigation, it is impossible to reach this goal, but to avoid more than one infection in the given situation should be aimed for. Equation (8) can therefore be transformed into equation (9) (individual risk) and (10) (number of persons getting infected in the considered situation).

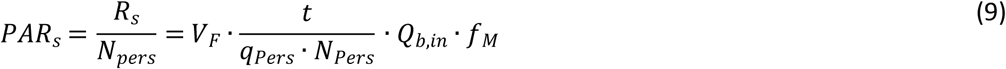

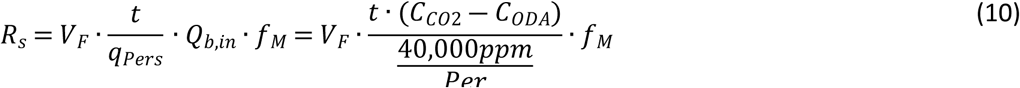

### 2. CO_2_-threshold for a certain infection risk

If R_s_ is limited to 1 to avoid a spread of the infection, the CO2 threshold in this situation can be calculated as:

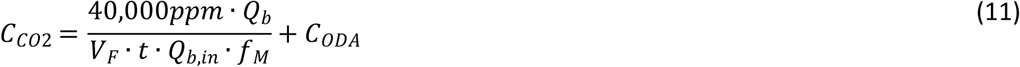

With the further assumption that the breathing activity of all persons in the room is the same it further simplifies into:

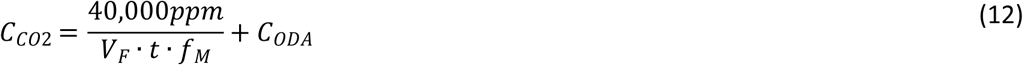

### 3. CO_2_-monitoring

If the CO_2_-concentration in the room is constantly monitored, the inhaled dose of CO_2_ (above outdoor concentration, *c*_*ODA*_) as well as the situational R-value can be calculated using equation (13). Instead of the integral, an approximation with the sum of the measured CO_2_-concentration can be used.

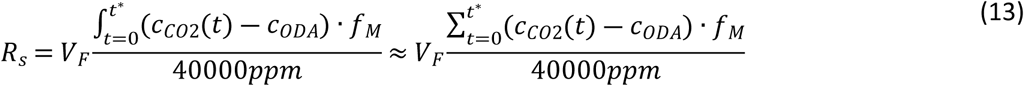

These equations can be used to set up tools for evaluating the infection risk in different situations.

## Results

For an exemplary viral emission of V_F_=47 1/h (mask efficiency f_M_=0.5 (surgical mask), outdoor air concentration 450 ppm), the different approaches should be applied. Figure 1 uses the mask type, the CO_2_-concentration and the time of stay as input parameters to calculate the risk of infection regarding equation (10). Besides measured CO_2_-concentrations, values for very good (600-800 ppm), good (900-1,100 ppm), moderate (1,400-1,600 ppm) and poor ventilation (1,900-2,100 ppm) are visualized. These ranges are based on the German national appendix of DIN EN 16798-1:2022-03 [15]. Three different mask types are visualized in Figure 1. No mask with a mask efficiency of f_M_=1.0, a surgical mask with a mask efficiency ranging from f_M_=0.3 to 0.7 (orange lines), but typical values of f_M_=0.4 to 0.6 (bar) and a FFP2 mask with a maximum range from f_M_=0.05 to 0.5 (lines), but a typical range of f_M_=0.12 to 0.20 (bar).

**Figure 1:**
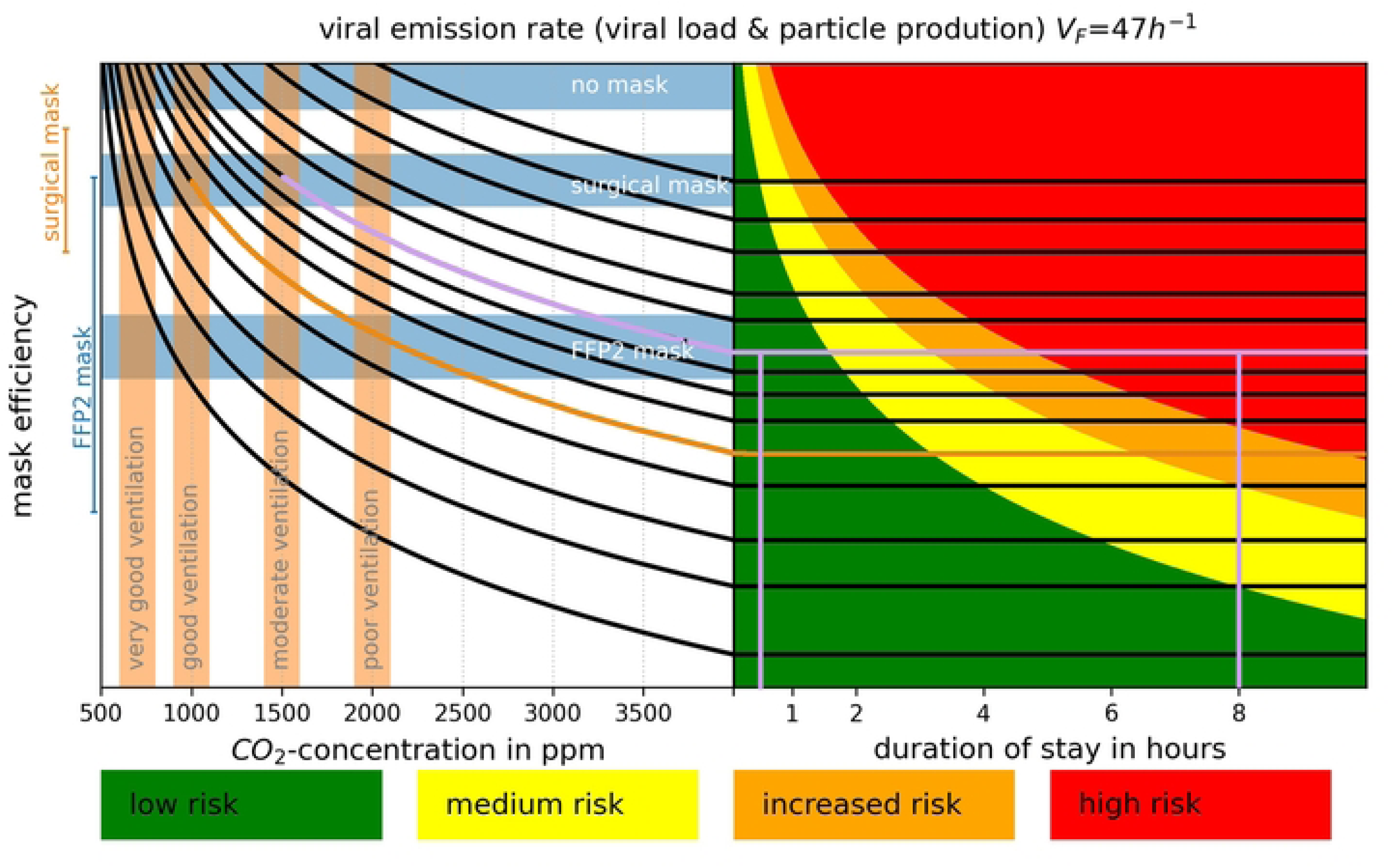
Prediction of the situational R-value for a viral emission rate (viral load & particle production) V_F_=47 h^-1^. Considers a quasi-steady CO_2_-concentration and a given mask-type for all attending persons. Very good ventilation (600-800 ppm), good ventilation (900-1,100 ppm), moderate ventilation (1,400-1,600 ppm), poor ventilation (1,900-2,100 ppm). No mask (f_M_=1,0), surgical mask (lines f_M_=0.3-0.7, bar f_M_= 0.4-0.6), FFP2-mask (lines f_M_=0.05-0.5, bar f_M_=0.12-0.2). Low Risk (green) for an R_s_<1, medium risk for 1<R_s_<2, increased risk for 2<R_s_<3 and high risk for R_s_>3

In the appendix of this study, further diagrams for V_F_=150 h^-1^ (Figure 5) and V_F_=500 h^-1^ (Figure 6) can be found. A low risk is seen if less than one person will probably get infected in this situation (green), an average risk for one to two persons (yellow), an increased risk for two to three persons (orange) and a high risk for more than three persons (red).

**Figure 2:**
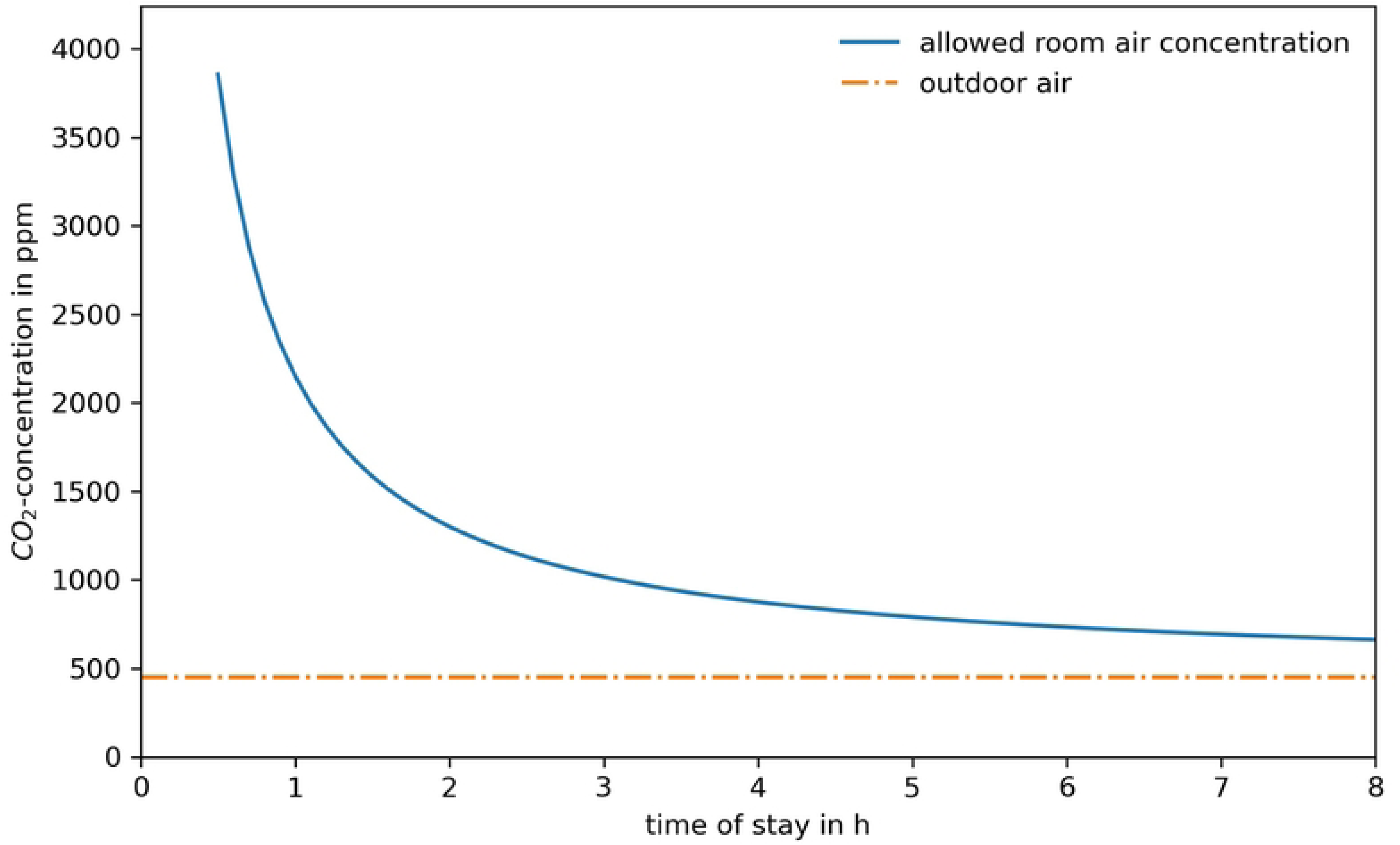
Allowed CO_2_-concentration in the room over time of stay for a viral emission of V_F_=47 1/h

**Figure 3:**
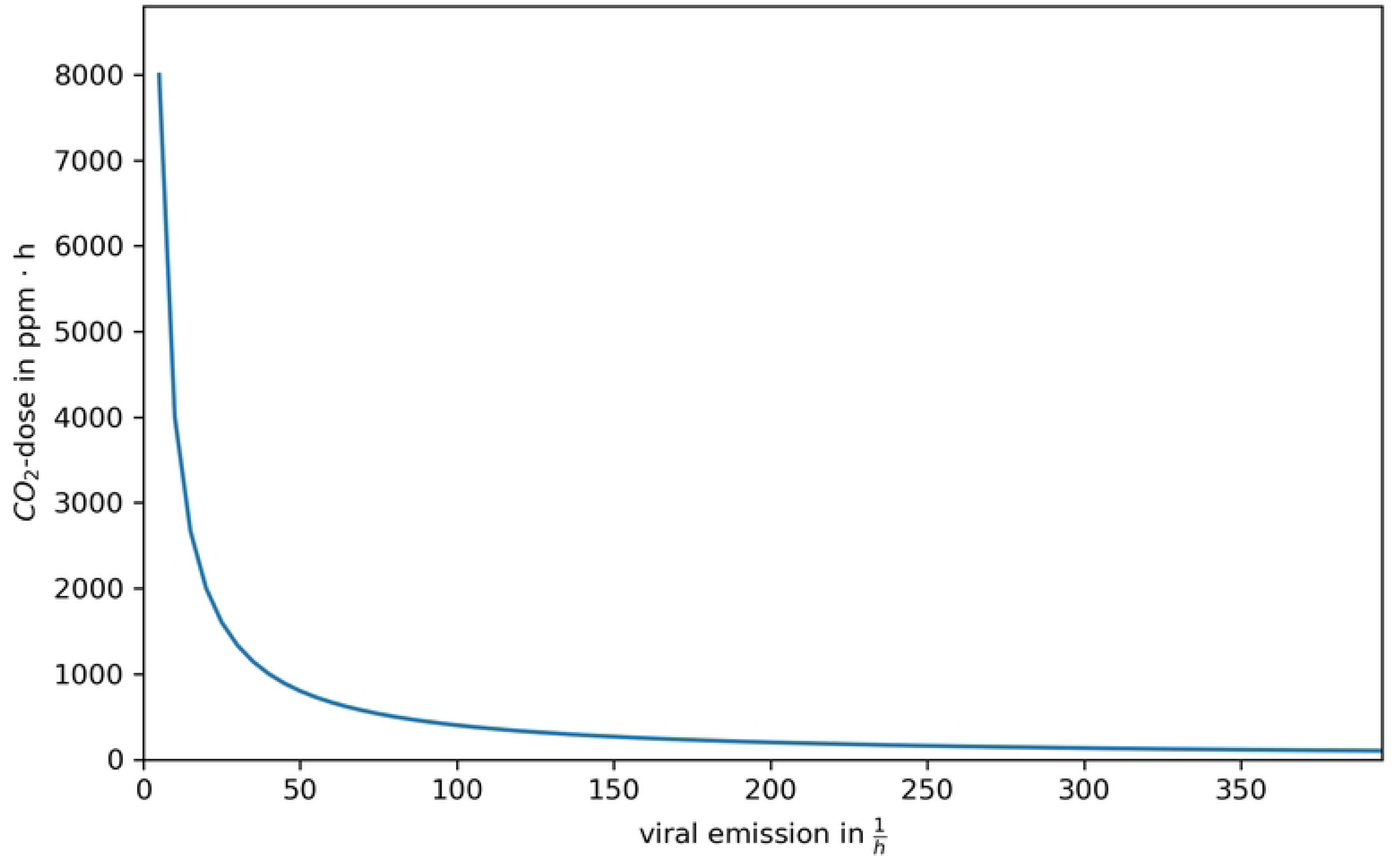
inhaled CO_2_-dose above outdoor level per hour of stay of the suspicious person depending on the viral emission of the infectious person without mask

**Figure 4:**
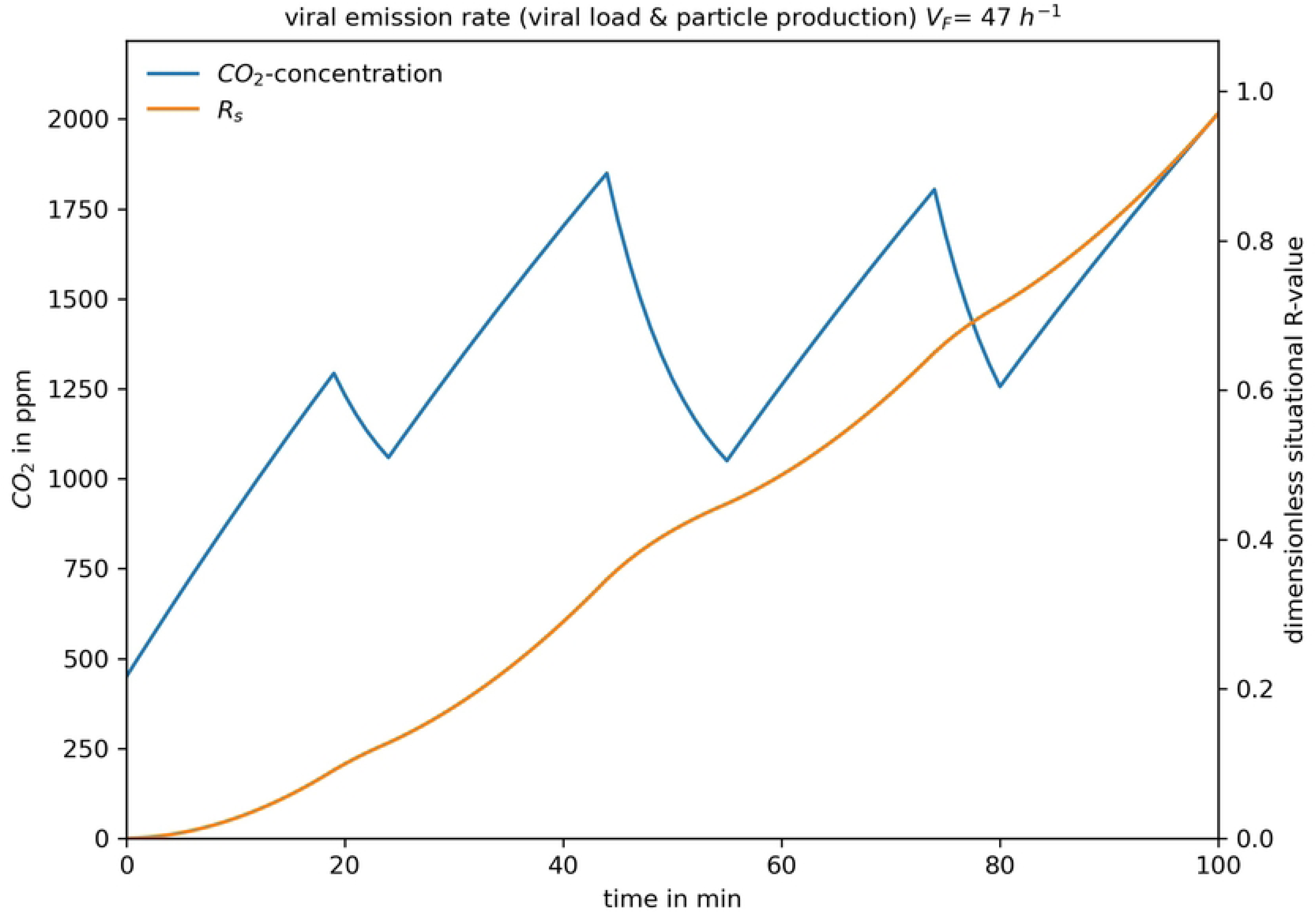
Exemplary Progress of the CO_2_-concentration during a school lesson (blue line) and resulting situational R-value over time (orange line)

**Figure 5:**
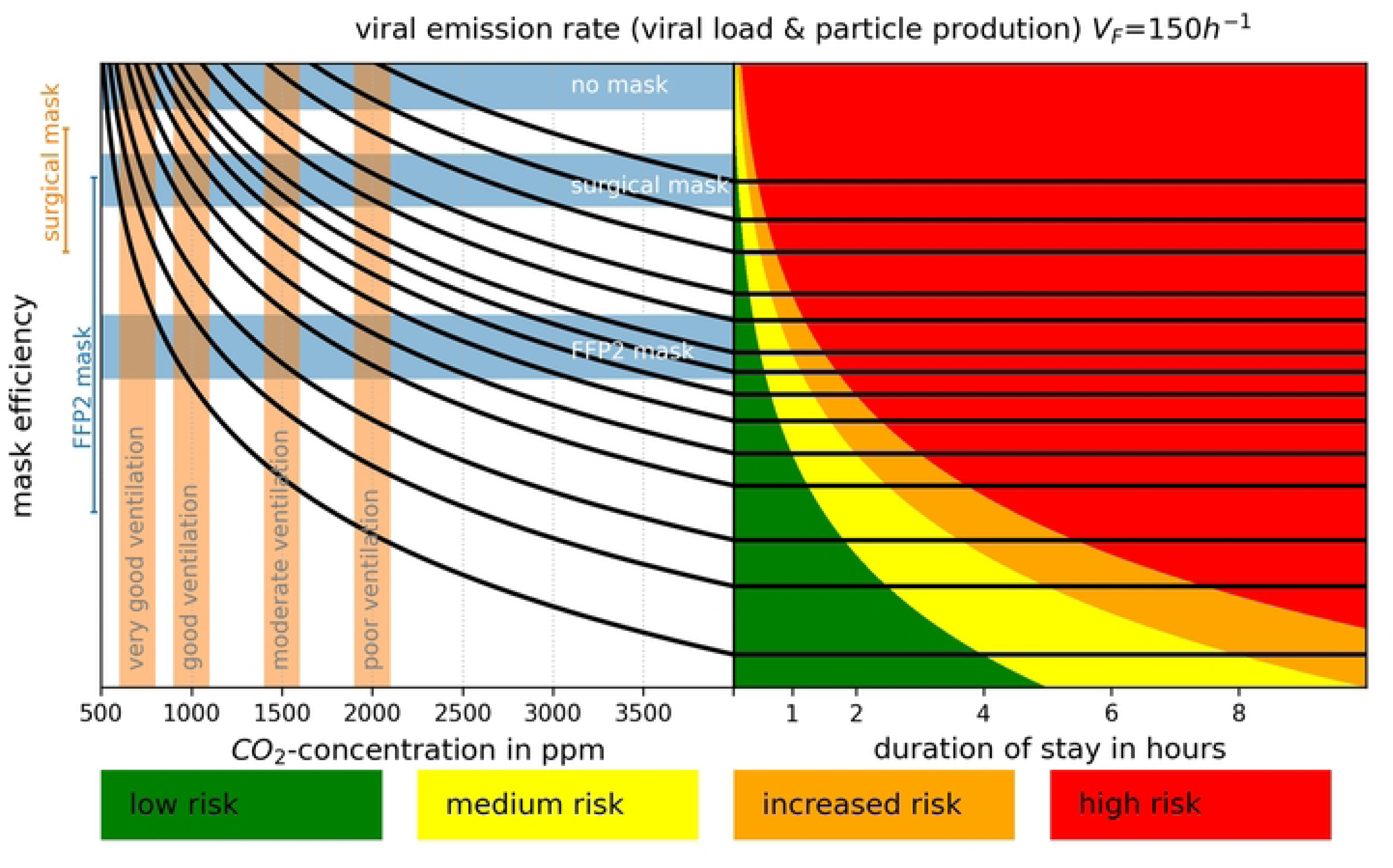
Prediction of the risk of infection for a viral emission rate (viral load & particle production) V_F_=150 h^-1^. Considering a quasit-steady CO_2_-concentration and a given mask-type for all attending persons. Very good ventilation (600-800 ppm), good ventilation (900-1,100 ppm), moderate ventilation (1,400-1,600 ppm), poor ventilation (1,900-2,100 ppm). No mask (f_M_=1,0), surgical mask (lines f_M_=0.3-0.7, bar f_M_= 0.4-0.6), FFP2-mask (lines f_M_=0.05-0.5, bar f_M_=0.12-0.2). Low Risk (green) for an R_s_<1, medium risk for 1<R_s_<2, increased risk for 2<R_s_<3 and high risk for R_s_>3

**Figure 6:**
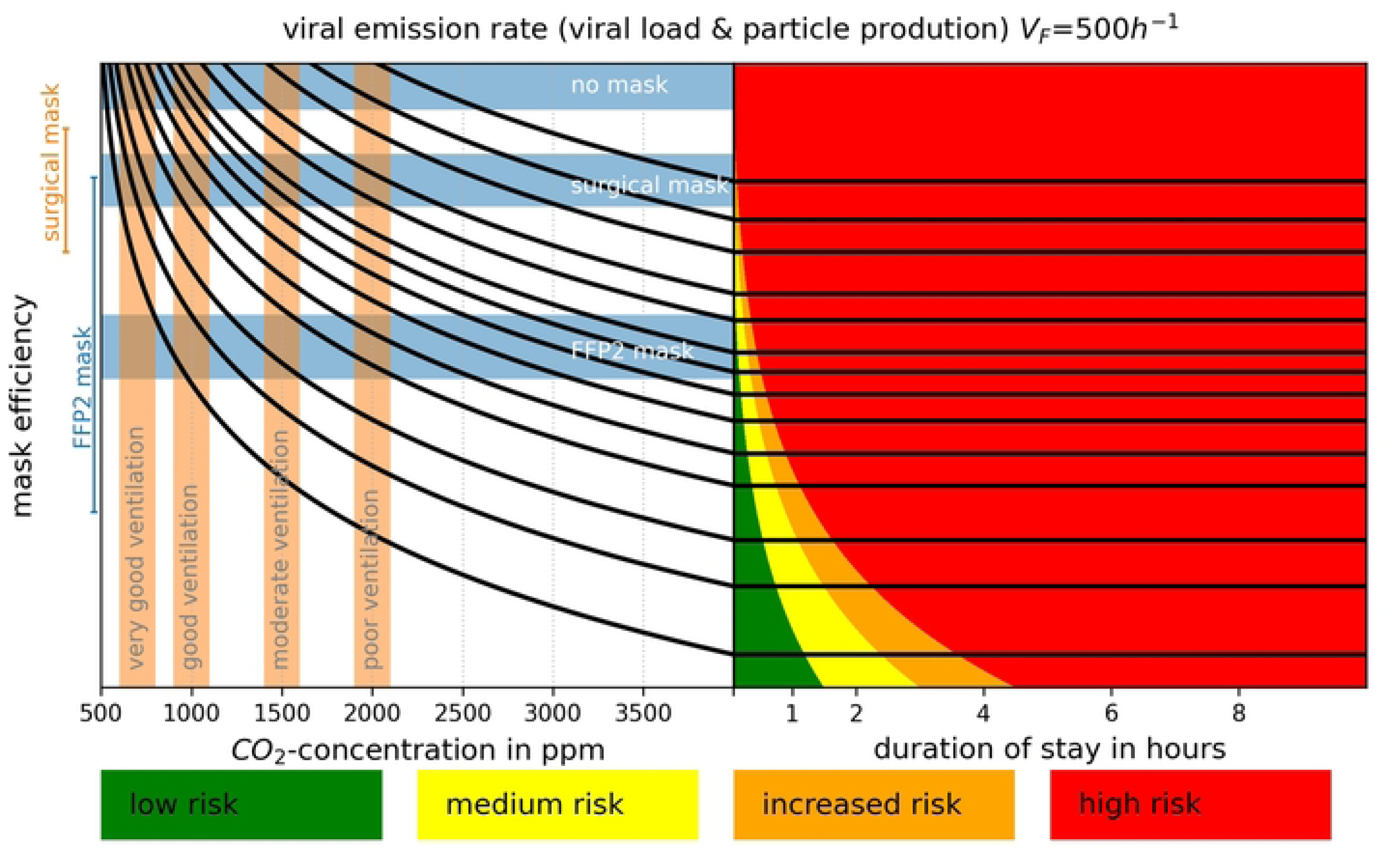
Prediction of the risk of infection for a viral emission rate (viral load & particle production) V_F_=500 h^-1^. Considers a quasit-steady CO_2_-concentration and a given mask-type for all attending persons. Very good ventilation (600-800 ppm), good ventilation (900-1,100 ppm), moderate ventilation (1,400-1,600 ppm), poor ventilation (1,900-2,100 ppm). No mask (f_M_=1,0), surgical mask (lines f_M_=0.3-0.7, bar f_M_= 0.4-0.6), FFP2-mask (lines f_M_=0.05-0.5, bar f_M_=0.12-0.2). Low Risk (green) for an R_s_<1, medium risk for 1<R_s_<2, increased risk for 2<R_s_<3 and high risk for R_s_>3

For the exemplary boundary conditions (f_M_=0.5, C_ODA_=450 ppm, C_CO2_=1500 ppm), the risk is low for a stay of 0.5 h, but high for 8 h (violet line). With these diagrams, different measures (e.g. change in CO_2_-concentration, mask type, time of stay) can easily be compared. So a decrease in the CO_2_-concentration by 500 ppm (good ventilation) with otherwise unchanged boundary conditions would result in a reduction of the risk for the long stay down to an increased risk (brown line), which could be further reduced by the use of an FFP2 mask.

Equation (12) can be used to calculate the maximum allowed (average) CO_2_-concentration. For a stay of half an hour, a CO_2_-concentration of approximately 3850 ppm can be allowed (see Figure 2), but for 8 h the allowed CO_2_-concentration (660 ppm) is just slightly above the outdoor concentration (450 ppm). For the longer stay heavy ventilation is necessary, which might be difficult to reach in uncomfortable outdoor conditions (cold, rainy, …) as well as unfavorable conditions (little wind, little temperature difference between indoor and outdoor). In these cases, further measures, e.g. better masks, testing, reducing number of persons, … have to be applied to meet the criteria of no more than one newly infected person after the considered situation.

From these calculations a critical CO_2_-dose above outdoor level can be found regarding equation (14), but it heavily depends on the viral emission of the infectious person as it can be seen in Figure 3. In Figure 3 no mask (f_M_=1) is considered, but it can be easily integrated by dividing the read value by the mask efficiency.

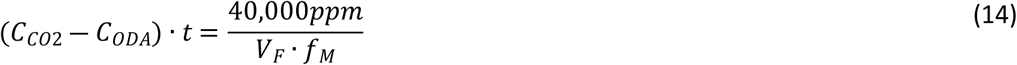

In Figure 4 the theoretical progress of the CO_2_-concentration in an exemplary classroom is displayed (blue line). A classroom with a volume of 150m^3^, 20 persons and window ventilation every 20 min was used to set up this theoretical progress of the CO_2_-concentration. The progress of the situational R-value is calculated regarding equation (13). After a double lesson of 2 × 45 min with a break of 10 min in between, one person would be infected in this example. A decrease of the CO_2_-concentration during window ventilation can also be seen in the course of the situational R-value as a flatter curve.

## Discussion/Limitations

The considerations presented in this paper are based on a model set up by Kriegel et. al. [1]. This model, like many comparable models, is based on the model by Riley [24] and Wells [25], but is applied to the current pandemic virus SARS-CoV-2. All these models improved during the course of the pandemic as more and more knowledge regarding virus specific parameters (e.g. viral emission rate, virus activity in air, …) was gained. Still, the models are mainly based on known outbreaks and therefore tend to overestimate the real risk, especially if mainly large outbreaks are documented as occurred in the beginning of the pandemic. But even if the calculated risk is somewhat higher than the real risk, a comparison of different situations or different intervention measures is possible with the use of these models. Furthermore, the assumptions were slightly corrected by medical investigations and may therefore be more accurate now.

This study now focusses on the implementation of the CO_2_-concentration into this model. Whereas the CO_2_-concentration within a room depends on the ventilation rate, the room size and the activity of the room users, it is a useful indicator for the ventilation rate in a steady state situation. For these calculations, a steady state situation is therefore assumed. This assumption is appropriate for long stays or small rooms, which are both cases with a potentially higher risk of infection, and therefore these cases which would more likely be considered by such models. Furthermore, for short stays or large rooms, the model may overestimate risk. In most cases the risk is not that high anyway so an overestimation is not considered critical, because it will probably not result in cost-intensive intervention measures which might not have been necessary.

In most cases, the exact viral emission rate is unknown and therefore the most import uncertainty within the method to set up non-medical interventions for different indoor environments. So the uncertainty of other parameters (e.g. the measured CO_2_-concentration in the room and outdoor) may also influence the outcome, but not as much as the viral emission rate itself. In addition, it is assumed that the air flow is the only mechanism that transports virus laden particles out of the room. In rooms with either appropriate working mechanical ventilation or regular window ventilation this might be reasonable, but it might be critical if an increased CO_2_-concentration is measured in the room. But once more this will overestimate the risk of infection in such critical cases. As it can be seen in Figure 1, without a FFP2 mask the risk in these cases will be evaluated as increased or high after a very short stay (not more than 30 min) even with an average viral emission rate, so an implementation of this discharge mode will increase the possible stay a little bit, but not significantly. For a first estimation of the effectiveness of different preventive measures, the use of the simplified model may therefore be appropriate.

Besides the aforementioned limitations regarding the simplifications of the model, the dynamics of the pandemic and especially the spread of new variants may heavily influence the input parameters of the model and therefore the significance of the results presented within this paper. Still the equations can easily be adapted to the new boundary conditions, if necessary.

## Conclusions

CO_2_ has been used as an indicator for indoor air quality for many years. In general, it is used as an instantaneous value and therefore reaches its limits if the dose is important, such as for the inhalation of potentially virus laden particles. Still, in this paper it is shown that continuously monitoring CO_2_ may be helpful to evaluate the amount of fresh air supplied to the room and therefore, besides improving the indoor air quality, will reduce the inhaled dose of potentially virus laden particles. The diagrams set up for different viral emission rates can be useful in comparing different non-medical measures and can easily be adapted to new variants or even new viruses if they are transmitted by aerosol particles as well, and the important values (e.g. viral emission rate) are known.

## Data Availability

All data presented in the paper.

## Figures in Appendix

## Literature

[1] M. Kriegel, A. Hartmann, U. Buchholz, J. Seifried, S. Baumgarte and P. Gastmeier, “SARS-CoV-2 Aerosol Transmission Indoors: A Closer Look at Viral Load, Infectivity, the Effectiveness of Preventive Measures and a Simple Approach for Practical Recommendations,” Preprint MedRxiv, DOI: https://doi.org/10.1101/2021.11.04.21265910, 2021.

[2] E. Ding, D. Zhang and P. M. Bluyssen, “Ventilation regimes of school classrooms against airborne transmission of infectious respiratory droplets: A review,” Build. Environ. 207, 108484, doi: https://doi.org/10.1016/j.buildenv.2021.108484, 2022.

[3] C. Sun and Z. Zhai, “The efficiacy of social distance and ventilation effectiveness in preventing COVID-19 transmission,” Sustain. Cities Soc. 62, doi: https://doi.org/10.1016/j.scs.2020.102390, 2020.

[4] L. Chatzidiakou, D. Mumovic and A. Summerfield, “Is CO2 a good proxy for indoor air quality in classrooms? Part 1.: The interrelationsships between thermal conditions, CO2 levels, ventilation rates and selected indoor pollutants,” Build. Sev. Eng. Res. Technol. 26, doi: https://doi.org/10.1177/0143624414566244, pp. 129–161, 2015.

[5] J. Bartyzel, D. Zięba, J. Nęcki and M. Zimnoch, “Assessment of Ventilation Efficiency in School Classrooms Based on Indoor-Outdoor Particle Matter and Carbon Dioxide Measurements,” Sustainability 12, 5600, doi: https://doi.org/10.3390/su12145600, 2020.

[6] S. Batterman, “Review and Extension of CO2-Based Methods to Determine Ventilation Rates with Application to School Classrooms,” Int. J. Environ. Res. Public Health, 14, 145, doi: https://doi.org/10.3390/ijerph14020145, 2017.

[7] A. Kabirikopaei and J. Lau, “Uncertainty analysis of various CO2-Based tracer-gas methods for estimating seasonal ventilation rates in classrooms with different mechanical systems,” Build. Environ. 179, 107003, doi: https://doi.org/10.1016/j.buildenv.2020.107003, 2020.

[8] L. Spinelle, M. Gerboles, M. G. Villani, M. Aleixandre and F. Bonavitacola, “Field calibration of a cluster of low-cost commercially available sensor for air quality monitoring. Part B: NO, CO and CO2,,” Sensors Actuators B Chem. 238, pp. 706–715, 2017.

[9] D. Olsson, “History of Ventilation Technology,” Swegon Air Acad., https://www.swegonairacademy.com/siteassets/_documents/history-of-ventilation-technology.pdf, 2015.

[10] W. G. Locher, “Max von Petternkofer (1818-1901) as a pioneer of modern hygiene and preventive medicine,” Eviron. Health Prev. Med. 12, doi: https://doi.org/10.1007/BF02898030, pp. 238–245, 2007.

[11] U. Satish, M. J. Mendell, K. Shekhar, T. Hotchi, D. Sullivan, S. Streufert and W. J. Fisk, “Is CO2 an Indoor Pollutant? Direct Effects of LOw-to-Moderate CO2 Concentration on Human DecisionMaking Performance,” Environ. Health Perspect. 120, doi: https://doi.org/10.1289/ehp.1104789, pp. 1671–1677, 2012.

[12] B. Du, M. C. Tandoc, M. L. Lack and J. A. Siegel, “Indoor CO” concentration and cognitive function: A critical review,” Indoor Air 20, doi: https://doi.org/10.1111/ina.12706, pp. 1067–1082, 2020.

[13] ASHRAE, “ASHRAE Standard 62.1-2019 Ventilation for Acceptable Indoor Air Quality,” https://ashrae.iwrapper.com/ViewOnline/Standard_62.1-2019, 2019.

[14] ISO, “ISO 17772-1:2017 Energy performance of buildings - Indoor Environmental Quality - Part 1: Infoor Environmental Input Parameters for the Design and Assessment of Energy Performance of Buildings,” 2017.

[15] ISO, “DIN EN ISO 16798-1: Energy performance of building - Ventilation for Buildings,” 2022.

[16] Ad-hoc-Arbeitsgruppe Innenraumrichtwerte der Innenraumlufthygiene-Kommision des Umweltbundesamtes und der Obersten Landesgesundheitsbehörden, “Gesundheitliche Bewertung von Kohlendioxid in der Innenraumluft,” Bekanntmachung des Umweltbundesamtes, doi: https://doi.org/10.1007/s00103-008-0707-2, 2008.

[17] Kommision Innenraumlufthygiene am Umweltbundesamt, “Das Risiko einer Übertragung von SARS-CoV-2 in Innenräumen lässt sich durch geeignete Lüftungsmaßnahmen reduzieren,” Umweltbundeamt - Für Mensch und Umwelt, 2020.

[18] J. Bhardwaj, S. Hong, J. Jang, C. H. Han, J. Lee and J. Jang, “Recent advancements in the measurement of pathogenic airborne viruses,” J. Hazard. Mater. 420, doi: https://doi.org/10.1016/j.hazmat.2021.126574, 2021.

[19] A. D. Gilio, J. Palmisani, M. Pulimeno, F. Cerino, M. Cacace, A. Miani and G. d. Gennaro, “CO2 concentration monitoring inside educational buildings as a strategic tool to reduce the risk of Sars-CoV-2 airborne transmission,” Environ. Res. 202, 101560, doi: https://doi.org/10.1016/j.envres.2021.111560, 2021.

[20] R. K. Bhagat, M. S. D. Wykes, S. B. Dalziel and P. F. Linden, “Effects of ventilation on the indoor spread of COVID-19,” J. Fluid Mech. 903, doi: https://doi.org/10.1017/jfm.2020.720, 2020.

[21] Z. Pang, P. Hu, X. Lu, Q. Wang, Z. O’Neill, M. J. Walker, M. O’Connor and A. McFerrin, “A smart CO”-based ventilation control framework to minimize the infection risk of COVID-19 in public buildings,” Build. Simul. 2021 Conf., https://www.researchgate.net/publication/349121056_A_Smart_CO2-Based_Ventilation_Control_Framework_to_Minimize_the_Infection_Risk_of_COVID-19_In_Public_Buildings, 2021.

[22] S. N. Rudnick and D. K. Milton, “Risk of indoor airborne infectionn transmission estiamted from carbon dioxide concentration,” Indoor Air 13, pp. 237–245, 2003.

[23] H. Dai and B. Zhao, “Association of the infection probability of COVID-19 with the ventilation rates in confined spaces,” Build. Simul. 13, doi: https://doi.org/10.1007/s12273-020-0703-5, 2020.

[24] E. Riley, G. Murphy and R. Riley, “Airborne Spread of Measles in a Suburban Elementary School,” American Journal of Epidemiology, vol. 107, no. 5, pp. 421–432, 1978.

[25] W. Wells, Airborne contagion and air hygiene: an ecological study of droplet infections, 1955.

[26] C. M. Issarow, N. Mulder and R. Wood, “Modelling the risk of airborne infectious disease using exhaled air,” J. Theor. Biol. 372, doi: https://doi.org/10.1016/j.jtbi.2015/02.010, 2015.

[27] A. Hartmann and M. Kriegel, “Risk assessment of aerosols loaded with virus based on CO2-concentration,” preprint deposit once, doi: https://doi.org/10.14279/depositonce-10362, 2020.

[28] H. C. Burridge, S. Fan, R. L. Jones, C. J. Noakes and P. F. Linden, “Predictive and retrospective modelling of airborne infection risk using monitored carbon monoxide,” Indoor Built Environ., doi:; https://doi.org/10.1177/1420326X211043564, 2021.

[29] Z. Peng and J. L. Jimenez, “Exhaled CO” as a COVID-19 Infection Risk Proxy for Different Indoor Environments and Activities,” Environ. Sci. Technol. Lett. 8, doi: https://doi.org/10.1021/acs.estlett.1c00183, 2021.

[30] DIN, “DIN EN 16798-3: 2017-11 Energy performance of buildings - Ventilation for buildings - Part 3: For non-residential buildings - Performance requirements for ventilation and room- conditioning systems,” 2017.

[31] Verein Deutscher Ingenieure, “VDI 3804:2009-03 Raumlufttechnik - Bürogebäude,” 2009.

[32] M. v. Pettenkofer, Ueber Luft in den Schulen und Ermittlung der Grenze zwischen guter und schlechter Zimmerluft, München, 1859.

[33] N. Good, K. M. Fedak, D. Goble, A. Keisling, C. L’Orange, E. Morton, R. Phillips, K. Tamer and J. Volckens, “Respiratory Aerosol Emissions from Vocalization: Age and Sex Differences Are Explained by Volume and Exhaled CO2,” Environmental Science & Technology, doi: https://doi.org/10.1021/acs.estlett.1c00760, 2021.

